# Polygenic Scores and Environmental Factors in Psychiatric Disorders: Gene–Environment Interaction Analyses Using the iPSYCH Study

**DOI:** 10.1101/2025.10.07.25337407

**Authors:** Tahereh Gholipourshahraki, Linda Ejlskov, Birgitte Dige Semark, Genona Torruella Maseras, Jette Steinbach, Ole Sahlholdt Hansen, Carsten Bøcker Pedersen, Cynthia M. Bulik, Michael Eriksen Benros, Preben Bo Mortensen, Esben Agerbo, Liselotte Vogdrup Petersen, Bjarni Jóhann Vilhjálmsson

## Abstract

**Background:** Psychiatric disorders represent a significant global health burden with complex etiologies involving both genetic and environmental factors. However, while the main effects of genetic and environmental factors are frequently studied, their interplay in shaping psychiatric disorder risk remains poorly understood. This study investigates the interaction between polygenic scores (PGS) as proxies for genetic risk and environmental factors in the risk of psychiatric disorders.

**Method:** This study utilized the iPSYCH case-cohort sample (n = 141,265). Environmental factors, including region, urbanicity, parental socioeconomic status, parental age, parental psychiatric history and early-life exposures: autoimmune disease, brain injury, and central nervous system (CNS) infections, were obtained from Danish nationwide registers. Logistic regression was used to examine the effects of targeted disorder-specific PGSs, environmental factors and their interactions on psychiatric disorders. Analyses were adjusted for age, sex and ancestral principal components to account for population stratification.

**Results:** Both PGS and environmental factors were associated with psychiatric disorders. We found limited evidence of gene–environment interactions across the investigated psychiatric disorders. Most interaction terms were small and not statistically significant, but a few remained significant. These included a smaller PGS association with attention deficit hyperactivity disorder in Southern Denmark compared with the Capital Region, a reduced PGS association with bipolar disorder in the medium and lowest parental income groups compared with the highest income group, and a weaker PGS association with major depressive disorder in individuals with parental psychiatric history compared with those without such history. In contrast, a larger PGS association with schizophrenia was observed in the paternal age group 31–35 years compared with the 26–30 years reference group, and a stronger PGS association with anorexia nervosa in North Denmark compared with the Capital Region.

**Conclusion:** Genetic liability for psychiatric disorders, captured through PGS, was associated with mental disorder risk across environmental contexts. Only a few gene–environment interactions were significant, and these were modest and mental disorder-specific. Overall, the findings highlight the difficulty of detecting robust gene–environment interactions and the need for studies of sufficient scale and statistical power to identify subtle gene–environment effects and improve understanding of psychiatric disorder etiology.

## Introduction

Psychiatric disorders are among the leading contributors to the global burden of disease ^1^ and it is estimated that one in eight individuals is affected by a psychiatric disorder ^2^. Unlike most other common and chronic conditions, many psychiatric disorders tend to emerge during adolescence or early adulthood, and they can persist throughout the life of an individual and impact their daily functioning and quality of life ^3^, and have considerable societal impact ^1, 4 5^.

The etiology of psychiatric disorders is complex, with both genetic and environmental factors contributing ^6^. Many of these factors are shared across different mental disorders. The heritability of psychiatric disorders is estimated to range from 37% for major depressive disorders (MDD) ^7^ to 65%-80% for schizophrenia (SCZ)^8, 9^ and about 70-90 % for bipolar disorders (BP) ^10 11^. Nevertheless, it is well established that the heritability of complex conditions such as psychiatric disorders is attributable to a large number of common genetic variants, each contributing a small effect to disease susceptibility. These findings have prompted the use of polygenic scores (PGS) to understand their genetics ^12^.

Apart from genetic factors, multiple environmental factors are known to be associated with psychiatric disorders. Environmental risk factors for psychiatric disorders include prenatal and postnatal risk factors such as infection ^13^, malnutrition^14^, low birth weight^15, 16^, family history of psychiatric disorders ^17^, childhood environment, including exposure to infections ^18, 19^, living and growing up in an urban environment ^20^ and stressful life events ^21-23^. In addition to individual history, low socioeconomic status of the family during childhood ^24-27^ is also associated with some psychiatric disorders.

Gene-environment interaction refers to the phenomenon whereby the effect of genetic risk on an outcome is conditional on specific environmental exposures ^28^. Specifically, the predictive accuracy of PGS is influenced by environmental factors ^29, 30^ and non-transmitted parental genetics (i.e., parental alleles not inherited by the child that can still shape the child’s environment) ^31^. There are multiple methodological approaches to studying gene-environment interplay, such as interaction terms in regression models using individual genetic variants ^32^, genome-wide environment interaction studies ^33^, and family-based designs that help control shared familial confounding ^34^. One such approach can be the use of PGSs ^35^. PGS-based models are particularly well-suited to psychiatric traits, where individual single-nucleotide polymorphism (SNP) effect sizes are typically small and unlikely to yield robust interaction signals in isolation. Kendler et al.^36^ showed that, among monozygotic twins, exposure to stressful life events can have substantial moderating effects on the genetic risk of MDD. Similar interactions between PGS and childhood trauma have been observed in other studies of MDD ^37, 38^, while another study showed no interaction ^39^. While the majority of interactions between PGS and the environment have focused on MDD, few studies have investigated other psychiatric disorders ^40-45^. Moreover, a comprehensive investigation of interactions between PGS and environmental factors across major psychiatric disorders and diverse environmental exposures is currently lacking.

The aim of the present study was to investigate the interrelations between PGS for psychiatric disorders (including attention deficit/hyperactivity disorder (ADHD), autism (ASD), anorexia nervosa (AN), SCZ, schizophrenia spectrum disorders (SCZSP), BP, and MDD, and environmental factors at both the individual and family levels to psychiatric disorder risk. In contrast to this approach, most existing studies rely on case-control or cohort designs where individuals with psychiatric disorders may be overrepresented, which could potentially lead to biased effect estimation. To mitigate this issue, we applied inverse probability weighting (IPW)^46^ to adjust for differential sampling probabilities and improve the validity of our interaction estimates. Building on these considerations, we examined the extent to which variation in PGS and environmental exposures is associated with the risk of psychiatric disorders. We also assessed whether the association between PGS and psychiatric disorder risk differs across levels of environmental exposure, indicating a potential gene–environment interaction. An overview of the study design, data sources, and analytic workflow is presented in Figure 1.

**Figure 1.**
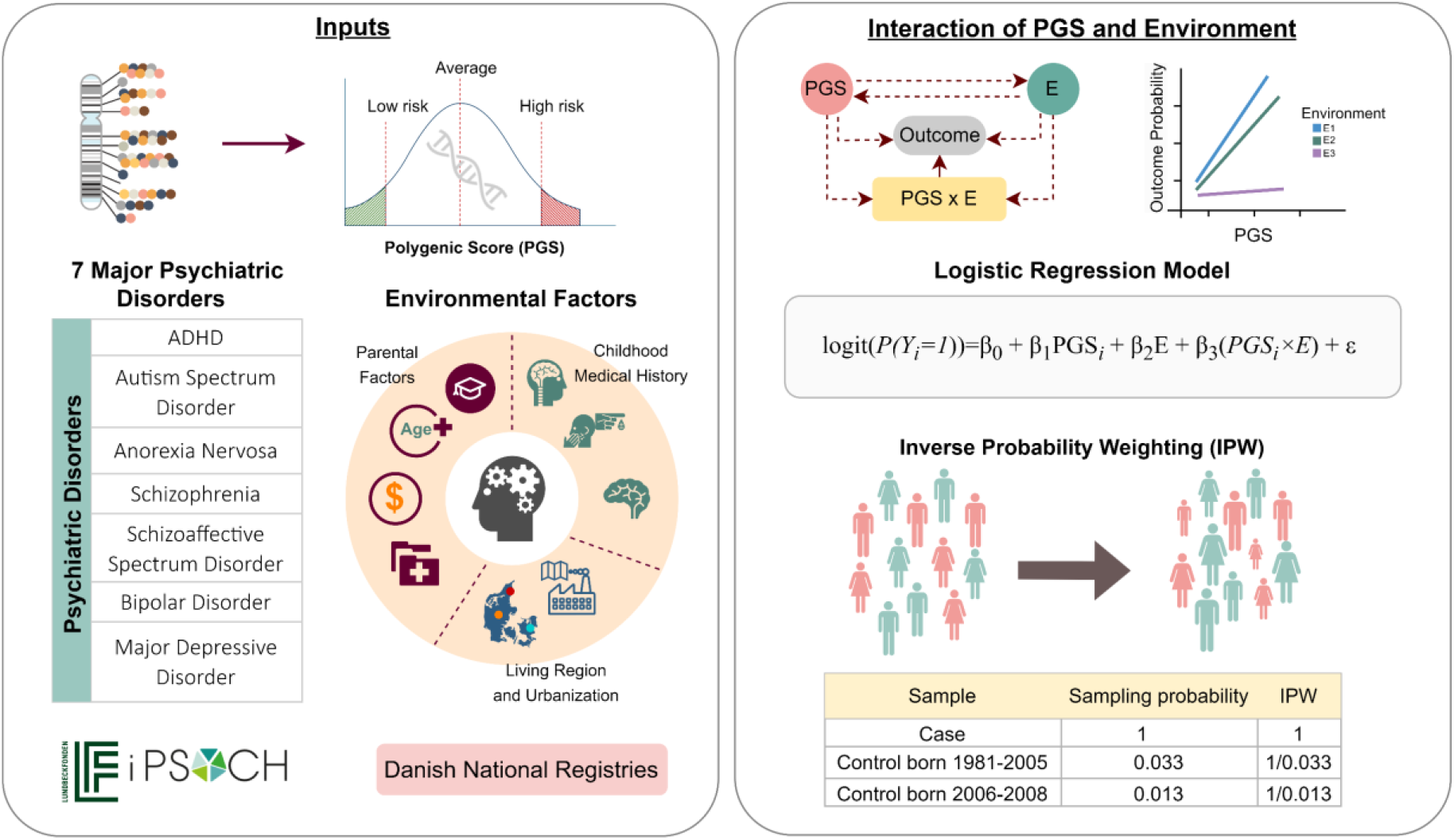
Overview of Study Design and Analytical Approach. This figure provides an overview of the study design, data sources, and analytical framework used to investigate gene-environment interactions in psychiatric disorders. The study analyzed seven major psychiatric disorders using data from the iPSYCH cohort, with polygenic scores (PGS) as genetic predictors and environmental factors obtained from Danish national registries. The interaction between PGS and environmental exposures was assessed using logistic regression models, incorporating inverse probability weighting (IPW) to account for sampling probabilities.

## Materials and Methods

### Study population

Participants were drawn from iPSYCH2015 (Lundbeck Foundation Initiative for Integrative Psychiatric Research), a nationwide, Danish case-cohort study of selected psychiatric disorders ^47^. We also included the AN samples from the Anorexia Nervosa Genetics Initiative (ANGI) ^48^, as they were samples within the same framework as iPSYCH2015. The iPSYCH2015 includes 141,265 singleton births in Denmark between 1981 and 2008 with known maternal information who were alive and living in Denmark at one year of age. The data includes 93,608 cases diagnosed with at least one of the major psychiatric disorders and a random population subcohort, including 50,615 controls. DNA extraction was based on the dried blood spot sample at birth, which has been stored in the Danish Neonatal Screening Biobank ^49^. Detailed information about the data can be found in the cohort’s documentation^50^.

All Danish residents are registered in the Danish Civil Registration System ^51^, which contains demographic information such as sex, date and place of birth, and parental identifiers. Each person is assigned a unique personal identification number, which enables accurate linkage of iPSYCH participants to nationwide health and social registers.

The iPSYCH study was approved by the Danish Scientific Ethics Committee, the Danish Health Data Authority, the Danish Data Protection Agency, Statistics Denmark, and the Danish Neonatal Screening Biobank Steering Committee. In accordance with Danish legislation, the Danish Scientific Ethics Committee waived the requirement for informed consent for this study, as it is based on existing biobank data.

### Measures

#### Exposures

Early life past medical histories: Data on a history of brain injury, central nervous system (CNS) infection before 10 years of age, and autoimmune diseases were obtained from the Danish National Patient Registries ^52^. Each category of medical history was analyzed separately. The histories were identified using International Classification of Diseases (ICD) codes from ICD-8 and ICD-10 (see Supplementary Table S1 for specific ICD codes).

Brain injury was characterized as any mild, moderate, or severe brain injury or skull fracture during childhood. Similarly, a binary indicator was developed for autoimmune disorders which included Type 1 diabetes, juvenile idiopathic arthritis (JIA), celiac disease, juvenile-onset systemic lupus erythematosus (SLE), rheumatic fever resulting in Sydenham chorea, and multiple sclerosis (MS).These conditions were selected as they represent some of the most common autoimmune disorders reported in previous studies to be associated with psychiatric disorders ^53-56^ and are characterized by onset in childhood, adolescence, or early adulthood, but rarely arise in late life. Lastly, a binary variable indicating any history of CNS infection during the first decade of life was identified as a potential risk factor.

Living region and urbanization: The Danish register data continuously updated residential address information since 1968. Since 2007, Denmark has been divided into 98 municipalities, each of which belongs to one of five regions which includes: North Denmark region, Central Denmark Region, Region of Southern Denmark, Capital Region, and Region Zealand. Data were collected on the location where a person had spent the most time during the first ten years of childhood, based on address records, and these addresses were merged with the corresponding region. Denmark was further subdivided into municipalities for the purpose of classifying the degree of urbanization ^57^ as: capital city, capital suburb, municipalities with more than 100000 inhabitants, municipalities with fewer than 100,000 inhabitants, and other municipalities in Denmark (largest town less than 10,000 inhabitants). As with region classification, each step of urbanization was capped at the individual’s primary residential address. The Capital Region, which has the highest population density and is the most urbanized region in Denmark, was chosen as the reference category.

Parental educational attainment (EA): The Danish education registries provided the information on the highest completed education for each parent ^58^. Educational qualifications were subdivided into three levels: mandatory education (Level 1); high school/vocational education (Level 2); and college or university (Level 3). The aggregate value of both parents’ education was classified into five categories from very low to very high.

Parental income: Annual income data of the mother and father were sourced from income registers. Related to this, some moderate year-to-year income fluctuations exist, including a negative tax-reported income in one year and a positive tax-reported income in the next. This is often the case for self-employed persons and business owners, but other reasons exist ^59^. To reduce the chances of misclassification, the five-year average for each parent’s income was computed at age 10 (meaning two years before and two years after the year the individual turned 10). Then, maternal and paternal income were summed to derive parental income. The total parental income was divided into quintiles, with quintile 1 as the lowest income group and quintile 5 as the highest.

Parental age at birth: Data on maternal and paternal ages at the time of birth were sourced from the Danish Central Population Register ^51^. Mother’s and fathers’ ages were analyzed separately and grouped into four categories: ≤25, 26–30 (reference), 31–35, and ≥36. The categorization was based on sample size distribution to avoid small groups (see Supplementary Table S2 for specific sample sizes by disorder).

Parental psychiatric history: A family history of psychiatric illness was defined by any recorded diagnosis of a psychiatric disorder (ICD F code) prior to the age of 10 years.

### Polygenic scores

The largest available GWAS summary statistics, excluding Danish samples, were used to calculate PGS for of ADHD ^60^, ASD ^61^, AN ^62^, SCZ and SCZSP ^63^, BP ^64^ and MDD ^65^. LDpred2-auto^66^ was used for PGS calculation. All analyses were statistically adjusted for 20 ancestral principal components (PCs) and the PGSs were z-standardized (mean = 0, SD = 1).

### Statistical analysis

#### PGS-environment interaction (PGSxE) analysis

To examine the association between PGS, environmental risk factors, and their potential interactions in relation to psychiatric disorder risk, we estimated a series of logistic regression models using the *glm2* package in R ^67^. First, we evaluated each PGS × environment interaction separately estimating an interaction term for each level independently. Second, we considered a model excluding interaction terms, to assess the extent to which the inclusion of interaction terms modified the estimated effects of PGS and environmental exposures. As the results obtained from these alternative specifications were consistent with those derived from a joint model, we presented the findings from the joint model only. This model simultaneously included the PGS, the environmental variable, and their interaction term. All models included age, sex, and PCs as covariates. Effects were reported as log-odds ratios (log OR) with 95% confidence intervals (CI), interaction terms were tested using Wald statistics, and multiple testing correction was applied using the Bonferroni method ^68^. All analyses were conducted in R version 4.4.1 ^69^. Detailed results are presented for ADHD, ASD, and MDD, which had the largest sample sizes, while results for the remaining disorders are provided in the Supplementary Tables (S3–S12) and Figures (S1-S4).

### Inclusion probabilities and weights

The iPSYCH2015 cohort was an expansion of the original study base, previously termed iPSYCH2012^70^. The source population of iPSYCH2012 consisted of individuals born in Denmark between May 1, 1981, and December 31, 2005, with 2.037% of the sample randomly selected as controls. In 2015, the cohort was expanded to include individuals born until December 31, 2008. This expansion incorporated a randomly selected control group with a probability of 1.267%, along with all cases diagnosed by the end of 2015.

Based on the sampling probabilities of the two iPSYCH subcohorts (2.037% and 1.267%), inclusion probabilities of 0.03278 and 0.01267 were assigned to subcohort members born in 1981–2005 and 2006–2008, respectively ^50^. IPW was applied in the model to account for sampling probabilities. Cases were assigned a weight of 1, while non-case subcohort members were assigned a weight equal to the inverse of their inclusion probability. This means that non-cases born in 1981–2005 were assigned a weight of 30.51 (= 1/0.03278), and those born in 2006–2008 were assigned a weight of 78.93 (= 1/0.01267). For individuals initially sampled as non-cases but later diagnosed as cases, the original IPW corresponding to their birth year was retained. IPWs were applied to account for sampling probabilities, and robust standard errors were used in all models to obtain valid inference under weighting.

## Results

### Sample characteristics

A summary of sample characteristics is presented in Table 1. The proportion of females was similar between the case and control groups (48.3% in total). The mean age at the time of analysis was 30.6 ± 7.02 years. The study included approximately 50,000 to 82,000 participants (depending on the disorder considered). A detailed description of the available data, including the respective numbers of environmental exposures, classification and additional categories, is provided in Supplementary Table S2.

**Table 1.** Summary statistics for key demographic and clinical characteristics in the iPSYCH2015 sample.

| Characteristic | Total (N = 134,603) | Cases (N = 89,628) | Controls (N = 44,975) |
| --- | --- | --- | --- |
| **Age\*** (mean ± SD) | 30.6 ± 7.02 | 31.04 ± 6.82 | 29.65 ± 7.31 |
| **Sex** (% Female) | 48,3% | 47.3% | 50.4% |
| **Number of cases/controls per disorder** |  |  |  |
| - ADHD | 73,235 | 27,543 | 45,692 |
| - Autism | 69,088 | 23,201 | 45,887 |
| - Anorexia nervosa | 53,470 | 7,126 | 46,344 |
| - Schizophrenia | 53,646 | 7,320 | 46,326 |
| - Schizophrenia Spectrum Disorders | 60,583 | 14,474 | 46,109 |
| - Bipolar Disorder | 49,925 | 3,461 | 46,464 |
| - Major Depressive Disorder | 82,218 | 36,811 | 45,407 |

ADHD: Attention-deficit hyperactivity disorder. Details on environmental exposures, including classification and additional categories, are provided in Supplementary Table S1.
\* Age at the time of analysis was calculated from participants' date of birth at the start of the study.

### Regional and urbanicity effects

The following results are based on joint models that included the PGS, the environmental variable, and their interaction term. In all models, the PGS effect corresponds to the reference category of the environmental exposure. Figure 2 displays log OR and 95% CI for the main PGS effects, environmental effects (region or urbanicity), and PGSxE terms. The results indicated that the risk of psychiatric disorders varies significantly across regions. Interestingly, a higher risk of ADHD was observed in Zealand and Central Denmark region compared to the Capital region. Similarly, living in the region of Central Denmark was associated with an increased risk of MDD.

**Figure 2.**
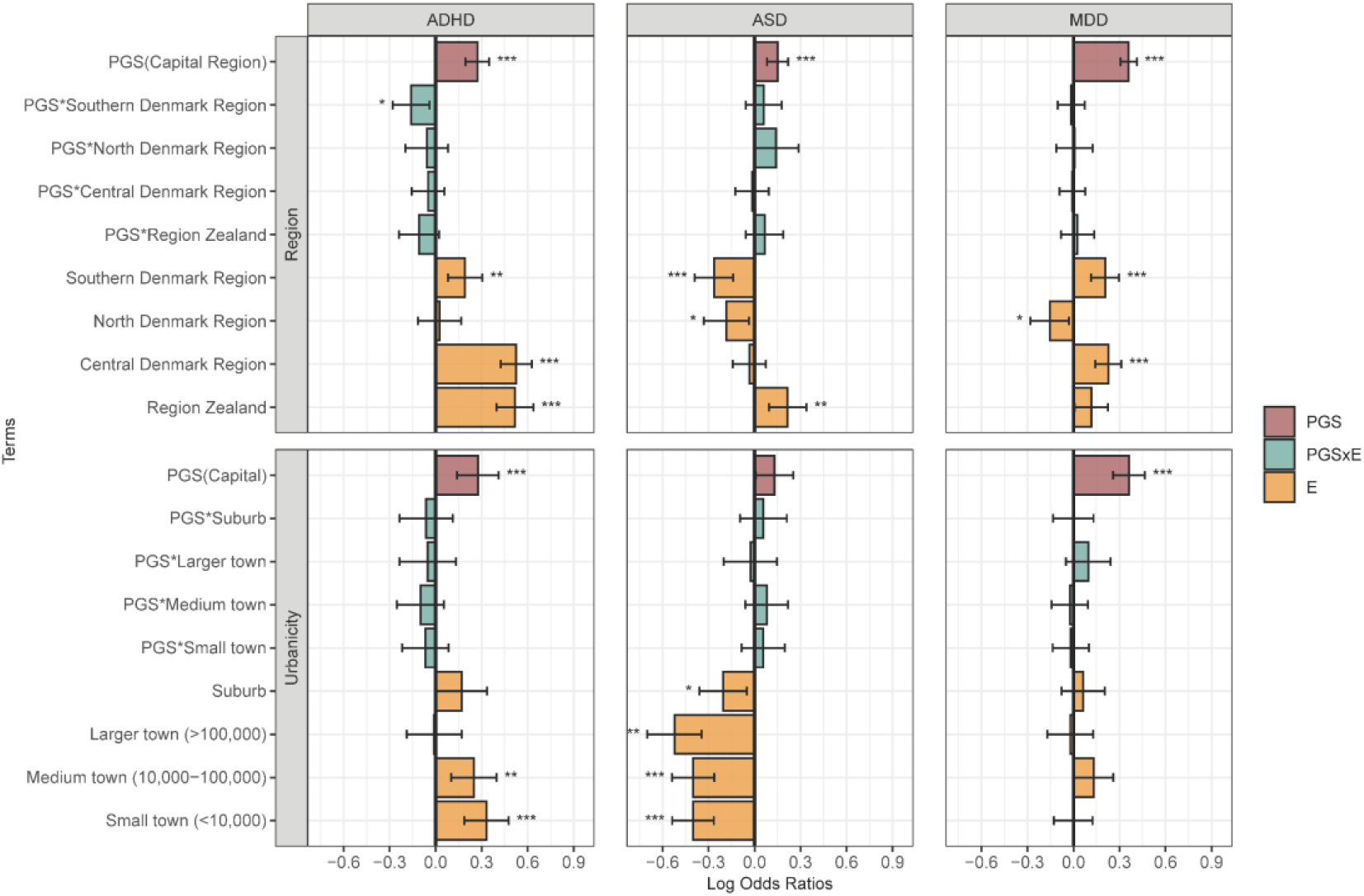
Region and urbanicity analysis. Log odds ratios and 95% confidence intervals are shown for each model term. The capital region serves as the reference category for both environmental variables. The PGS term represents the estimated effect of the polygenic score on the outcome within this reference category. The PGS×E (interaction) and E (environmental main effect) terms are interpreted relative to the reference category. Asterisks denote statistical significance at *p* < 0.05 *(), p < 0*.*01 (), and p < 0*.*001 ()*. ADHD: attention-deficit/hyperactivity disorder; ASD: Autism: autism spectrum disorder; MDD; major depressive disorder.

A few PGS×region interaction reached statistical significance. These included attenuation of PGS effects for ADHD in the Southern Denmark region (Figure 2) and stronger PGS effects for AN in the North Denmark region (Figure S1), compared with the Capital Region (reference category). For most other disorders, the PGS×region interactions were not statistically significant.

Differences in psychiatric disorder risk were also observed across levels of childhood urbanicity compared with the Capital Region. For instance, residence in less urbanized areas was associated with a lower risk of ASD. Similar patterns were observed for small towns (<100,000) in AN, SCZ, and schizophrenia spectrum disorders (Figure S1). No statistically significant PGS×urbanicity interactions were detected.

### Parental factors

Associations between parental characteristics, genetic risk, and their interactions were evaluated using joint models. The highest EA Category was used as the reference group. A significant direct effect of parental EA on all investigated psychiatric disorders was observed (see Figure 3 and Figure S2). In most disorders, lower EA levels were associated with an increased risk of psychiatric disorders compared to higher EA, except for AN, where a negative association was found (Figure S2). Interaction estimates were generally small and not statistically significant, which means the strength of PGS associations did not differ across education categories relative to the highest group.

**Figure 3.**
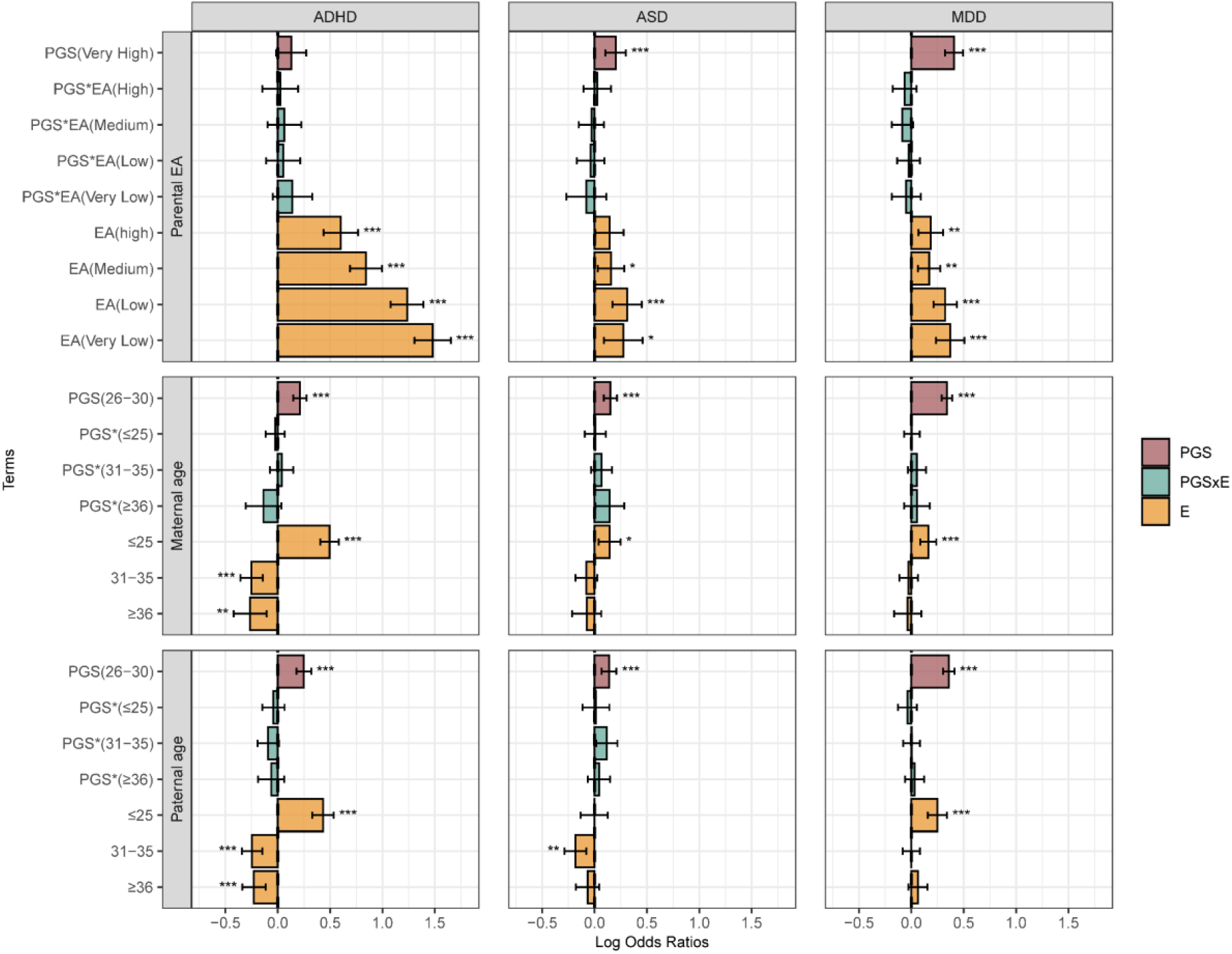
Parental education and parental age analysis. Log odds ratios and 95% confidence intervals are shown for each model term. The top panel includes parental educational attainment (EA), the middle panel includes maternal age, and the bottom panel includes paternal age. For parental EA, the Very High category serves as the reference group. For maternal and paternal age, the 26–30 category serves as the reference. The PGS term represents the estimated effect of the polygenic score on the outcome within this reference category. The PGS×E (interaction) and E (environmental main effect) terms are interpreted relative to the reference category. Asterisks denote statistical significance at *p* < 0.05 *(), p < 0*.*01 (), and p < 0*.*001 ()*. ADHD: attention-deficit/hyperactivity disorder; ASD: Autism: autism spectrum disorder; MDD; major depressive disorder.

Parental income showed a similar pattern, with lower income groups associated with increased risk compared with higher income (Figure S3). Two interactions remained statistically significant: the effect of the BP PGS was attenuated in the medium and lowest income groups compared with the highest income group. For other disorders, PGS×income estimates did not reach statistical significance.

Parental age at birth was analyzed separately for mothers and fathers, with the 26–30 years category used as the reference. A higher association with the risk of psychiatric disorders was observed for parental age ≤25 years for most disorders compared with the reference group. In contrast, statistically significantly smaller associations with the risk of ADHD were found for both maternal and paternal age. For most disorders, no clear evidence of modification of PGS associations was found. An exception was SCZ, where the PGS effect was stronger in the paternal age group 31–35 years compared with the reference (Figure S3).

A strong association was observed between parental psychiatric history and risk of psychiatric disorders, with higher risk among individuals with an affected parent compared with those without such history (Figure S4). One interaction reached statistical significance: for MDD, the PGS association with disorder risk was smaller in individuals with parental psychiatric history compared with those without. For the remaining disorders, the PGS×parental psychiatric history estimates were not statistically significant.

### Early life events

Figure 4 illustrates the relationship between early life exposures, including autoimmune disease history, childhood brain injury, and childhood CNS infection, and PGS.

**Figure 4.**
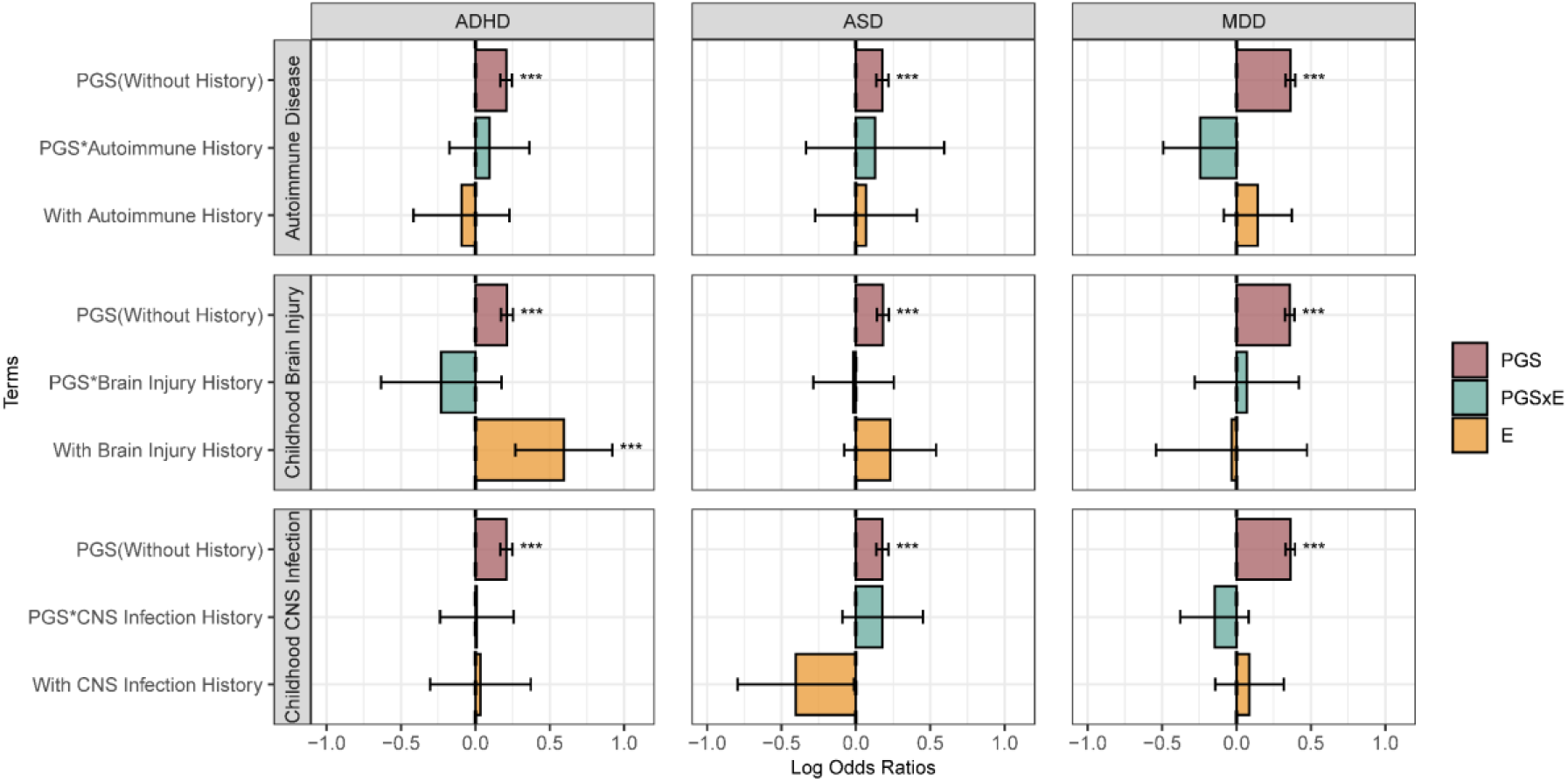
Early-life exposures analysis. Log odds ratios and 95% confidence intervals are shown for each model term. Panels represent models for (from top to bottom) autoimmune disease history, childhood brain injury, and childhood CNS infection. For each exposure, individuals without a history of the condition serve as the reference group. The PGS term represents the estimated effect of the polygenic score on the outcome within this reference category. The PGS×E (interaction) and E (environmental main effect) terms are interpreted relative to the reference category. Asterisks denote statistical significance at *p* < 0.05 *(), p < 0*.*01 (), and p < 0*.*001 ()*. ADHD: attention-deficit/hyperactivity disorder; ASD: Autism: autism spectrum disorder; MDD; major depressive disorder.

Among all exposures, only childhood brain injury showed a clear association with ADHD. For other disorders, no statistically significant associations with early-life histories were observed, which may be explained by the limited number of individuals with these exposures. The associations between PGS and psychiatric disorders did not differ from those in the unexposed reference groups.

## Discussion

The present study is, to the best of our knowledge, the most comprehensive investigation of the interaction between polygenic liability across the seven major psychiatric disorders and a wide range of early-life and contextual environmental factors. Using the largest population-based case-cohort dataset for studying mental health, and consistent modeling across outcomes, we found little evidence for robust PGSxE interactions. A small number of significant interactions were identified, but their effect sizes were generally small. Moreover, the direction and strength of these interactions varied, and no consistent pattern was observed across disorders or exposures. We also found that both PGSs and the environmental exposures considered were independently associated with the risk of psychiatric disorders.

We identified only a few statistically significant PGS×E interactions after accounting for direct genetic and environmental effects. These included attenuation of PGS effects for ADHD in Southern Denmark, for BP disorder in medium and low parental income groups, and for MDD in individuals with parental psychiatric history, compared with their corresponding reference categories. In contrast, stronger PGS effects were observed for SCZ in the paternal age group 31–35 years and for AN in North Denmark. For most other exposures, interaction estimates were small and not statistically significant, and no consistent cross-disorder pattern was observed.

However, the interpretation of a statistically significant PGSxE effect is difficult in practice, as it is affected by the choice of model used, the measuring scale for the environment (and outcome), genotype-variance effects, as well as other confounders. In general, it is difficult to rule out all of these possible confounders, especially if the effects are generally weak as in our case. The absence of a uniform trend may indicate that the mechanisms underlying gene-environment interactions are complex and disorder-specific. Furthermore, when considering interactions, a common assumption is that genetic susceptibility and environmental factors influence the same or related biological pathways, potentially modifying the outcome. Nevertheless, the mechanisms may be more intricate. As discussed by Westerman et al. ^71^, additional phenomena may contribute to these interactions. For instance, both genetic and environmental factors might influence a shared pathway or a specific mediator with a nonlinear relationship to the outcome. Moreover, their independent effects may exhibit nonlinear associations with the outcome. This may explain findings where the association between a specific environmental category and the outcome is stronger than its association with PGS when compared to the reference category. While PGSs are a useful tool for summarizing genetic liability, they may obscure locus-specific interactions. Genome-wide environment interaction studies could, in principle, identify variant-level interactions ^32^. Still, these approaches require considerably larger sample sizes than the >130.000 available in this study, particularly in the context of psychiatric disorders, which are highly polygenic. We should also note that the absence of a detected interaction effect between multiple environmental measures and PGS does not imply that gene–environment interactions do not exist. Instead, interaction effects may still occur at a more detailed or specific level, such as within particular subpopulations, at the level of individual genetic variants, or through more nuanced biological mechanisms that are not captured by broad pollution measures or aggregated PGSs.

Some of the identified environmental associations were strong and could not be easily explained by genetics, e.g., we identified large regional differences in mental disorder prevalences in Denmark, broadening the findings of a recent study displaying considerable geographical variation for ADHD and ASD in Denmark ^72^. These findings may suggest that regional factors, such as regional differences in diagnosis and healthcare access, lifestyle, or environmental exposures may contribute to psychiatric disorder risk. Related to this, urbanicity was also associated with differences in psychiatric risk. Living in less urbanized areas was associated with a lower risk of ASD compared to highly urbanized areas. Vassos et al. reported that, for most psychiatric disorders, individuals born in the capital had a higher incidence than those born in rural areas in Denmark ^20^.

Parental EA and income were particularly strong risk factors for developing psychiatric disorders, with lower socioeconomic status consistently linked to increased risk across multiple disorders. Noteworthy, several of the environmental associations for AN differed from this general pattern. Higher parental education and income were associated with increased risk of AN. These observations are consistent with previous Danish register-based studies reporting that AN risk is elevated in families with higher socioeconomic position for AN compared with other eating disorders ^73^. In addition, PGS for AN has been linked to higher cognitive performance in childhood, which may suggest the notion that the etiological profile of AN differs from that of other psychiatric disorders. A history of early-life autoimmune disease and childhood brain injury was also associated with higher risk for some disorders. These findings align with previous research indicating that early-life adversity, health-related exposures, and socioeconomic context play a role in psychiatric disorder risk, independent of genetic liability ^27, 74-78^.

Parental age at birth was also associated with psychiatric disorder risk. Younger maternal and paternal age were consistently linked to an increased risk across multiple disorders, a finding that is consistent with previous research suggesting that younger parental age may be associated with increased environmental stressors, less stable household environments, or genetic factors related to early reproduction ^79-81^. Interestingly, we did not find strong evidence for association between older parents and risk for ASD and other psychiatric disorders, which has been reported in other studies ^80-83^. However, as the analysis is restricted to the iPSYCH sample, it only includes parents born after 1980, who are very likely too young to observe the association between parental age and ASD among older parents.

The present study has several strengths. To our knowledge, this is the first study that utilized a large, population-based sample covering multiple psychiatric disorders, combined with comprehensive, nationwide register-based data on environmental exposures. In addition, we applied an IPW approach to account for sampling bias. This method enables more unbiased effect estimates and yields results that more closely approximate those we would expect if data from the entire underlying population were available. However, the interpretation of the results should be considered in light of several potential limitations. PGS estimates are derived from GWAS summary statistics, which may not fully capture rare variants, gene-gene interactions, or epigenetic modifications. PGSs also aggregate many genetic variants into a single score, which may dilute locus-specific effects and make it difficult to detect more nuanced gene–environment interactions. The predictive performance of PGS is further dependent on the discovery GWAS, and continued increases in sample size are expected to enhance the precision and utility of PGS. The limited number of individuals in certain environmental exposure categories (e.g., in the older parent age group) restricted further stratification of these effects. Furthermore, despite the large sample sizes, a few cases might exist in some specific category, and it is still possible that we were underpowered to detect an interaction effect. Another limitation arises from the time-varying variables. Risk factors might change over time, and the duration of risk factors is also relevant. The effect of each environmental factor may also depend on the specific disorder, such as whether it manifests in childhood or adulthood. Finally, misclassification of variables is possible, particularly for historical exposures recorded in national registers.

In conclusion, the results of this study suggest that PGS and both individual and prenatal factors are independent predictors of psychiatric disorders. We found little evidence for robust gene–environment interactions, as interaction estimates were generally small and not statistically significant. Importantly, there did not appear to be a unified pattern of environmental associations across disorders. Some disorders, such as AN, even showed environmental effects in the opposite direction compared with the other psychiatric disorders, which complicates the interpretation of PGS×E findings and emphasize the disorder-specific nature of these effects. Future research should aim to replicate these findings in diverse populations, explore additional environmental factors, and incorporate longitudinal designs to assess the timing of environmental exposures. Improved modeling techniques, such as integrating whole-genome risk scores or epigenetic data, could provide a more comprehensive understanding of gene-environment interplay. A deeper understanding of these complex relationships has the potential to inform context-aware approaches to risk assessment and early intervention in psychiatric disorders.

## Data Availability

The iPSYCH data used in this study are not publicly available due to legal and ethical restrictions. Researchers interested in accessing these data for research purposes may contact the iPSYCH Steering Committee and are subject to approval in accordance with iPSYCH data access policies.

