## Supplementory Table 1 for "Polygenic Scores and Environmental Factors in Psychiatric Disorders: Gene–Environment Interaction Analyses Using the iPSYCH Study"

Table S1. ICD-8 and ICD-10 codes for past medical history

| History category | ICD-8 Codes | ICD-10 codes |
| --- | --- | --- |
| Autoimmune disease | 249,712.0, 269.00, 734.10, 390, 392, 340 | E10, M08, M09, K900, M32, I00, I01, I02, G35 |
| Brain Injury | 850.99, 851.29-854.99, 800.99-801.09, 803.99 | S06.1-S06.9, S02.0, S02.1, S02.7, S02.9 |
| CNS infection | 013, 027.01, 036.09, 090.49, 094.9, 320.09-320.80, 322, 324, 392, 040-043.99, 045-046, 052.01, 053.02, 054.03, 055.01, 056.01, 062-065, 071.99, 072.02, 075.01, 079.29, 474, 320.89-320.99, 321, 323 | A17, A321, A390, A504, A521-A523, A692D, E236A, G00-G01, G042, G050, G060A-F, G061A-C, G062, G079A-B, I02, A80-A89, B003-B004, B010-B011, B020-B021, B050-B051, B060, B261-B262, G020, G051, P352A, A066, B375, B451, B582, G021-G028, G03, G040-G041, G048-G049, G052-G058, G060G-L, G061E-G, G079J-K, G08, G09 |
