## Supplementory Figure 1-4 for "Polygenic Scores and Environmental Factors in Psychiatric Disorders: Gene–Environment Interaction Analyses Using the iPSYCH Study"

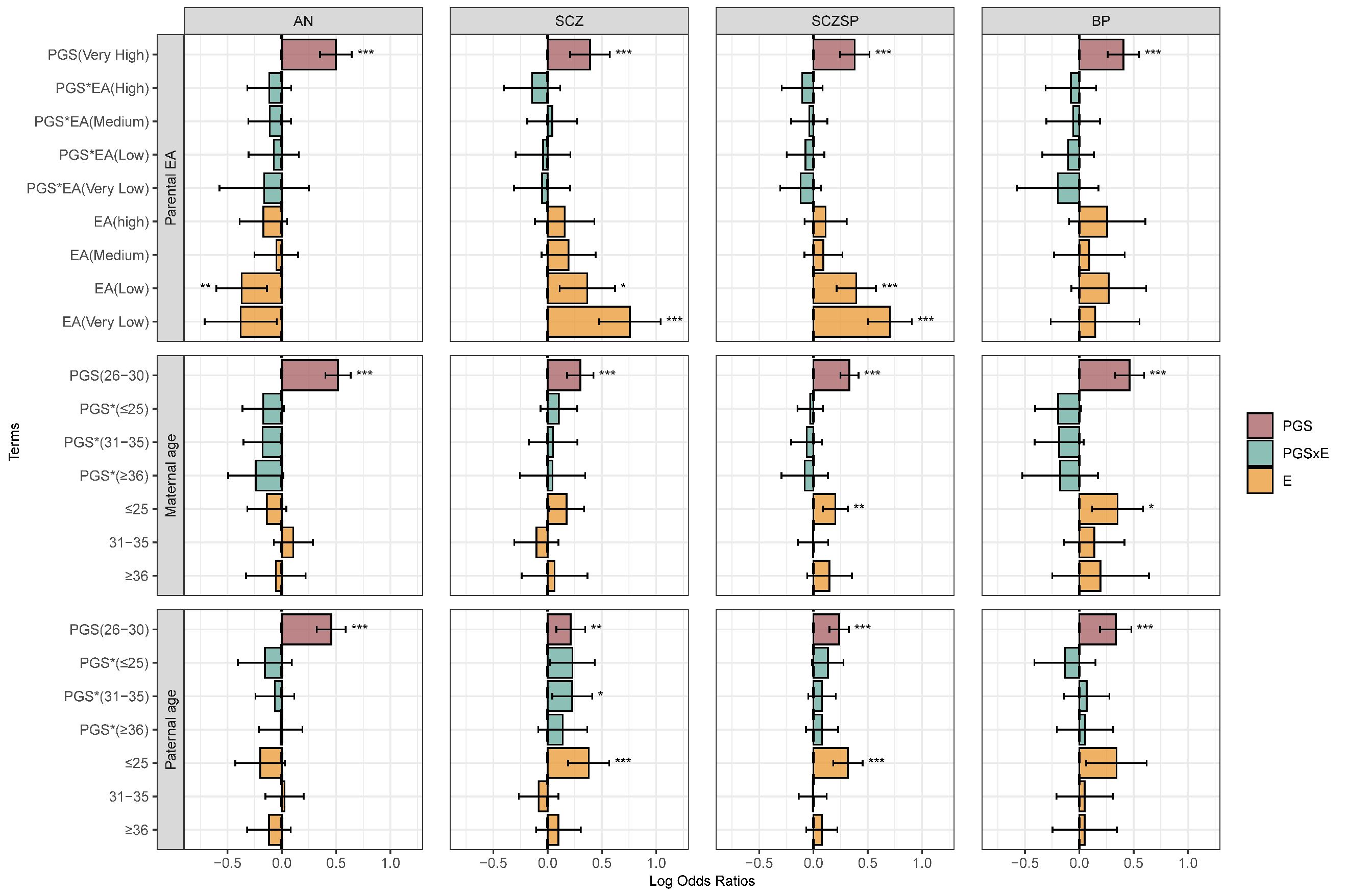

**S2 Figure. Parental education and parental age analysis.** Log odds ratios and 95% confidence intervals are shown for each model term. The top panel includes parental educational attainment (EA), the middle panel includes maternal age, and the bottom panel includes paternal age. For parental EA, the **Very High** category serves as the reference group. For maternal and paternal age, the **26–30** category serves as the reference. The **PGS** term represents the estimated effect of the polygenic score on the outcome within this reference category. The **PGS×E** (interaction) and E (environmental main effect) terms are interpreted relative to the reference category. Asterisks denote statistical significance at *p* < 0.05 *(), p < 0.01 (), and p < 0.001 ()*. AN: anorexia nervosa; SCZ: schizophrenia; SCZSP: Schizophrenia spectrum disorder; BP: Bipolar disorder.

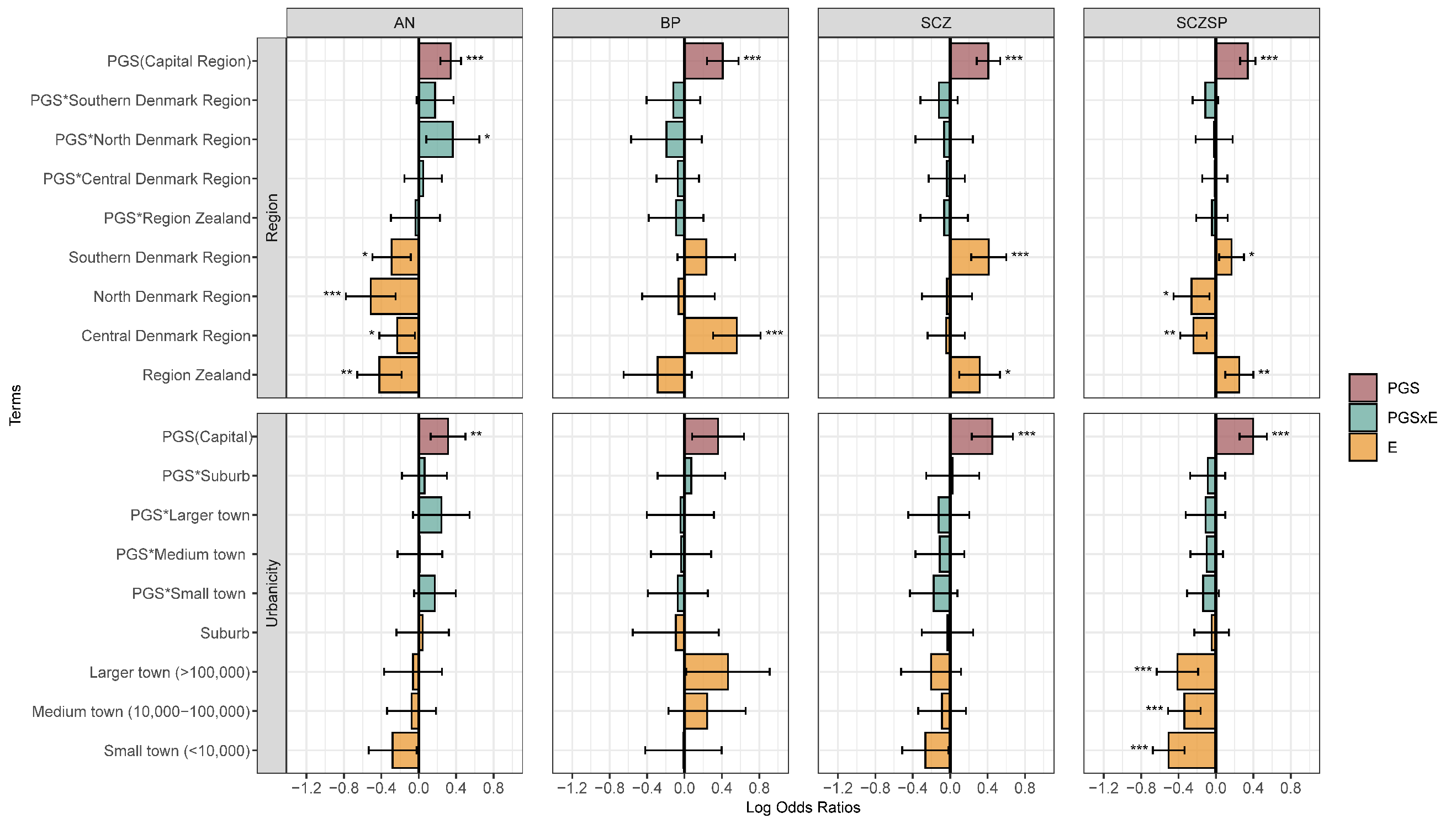

**S1 Figure. Region and urbanicity analysis.** Log odds ratios and 95% confidence intervals are shown for each model term. The capital region serves as the reference category for both environmental variables. The **PGS** term represents the estimated effect of the polygenic score on the outcome within this reference category. The **PGS×E** (interaction) and E (environmental main effect) terms are interpreted relative to the reference category. Asterisks denote statistical significance at *p* < 0.05 *(), p < 0.01 (), and p < 0.001 ()*. AN: anorexia nervosa; SCZ: schizophrenia; SCZSP: Schizophrenia spectrum disorder; BP: Bipolar disorder.

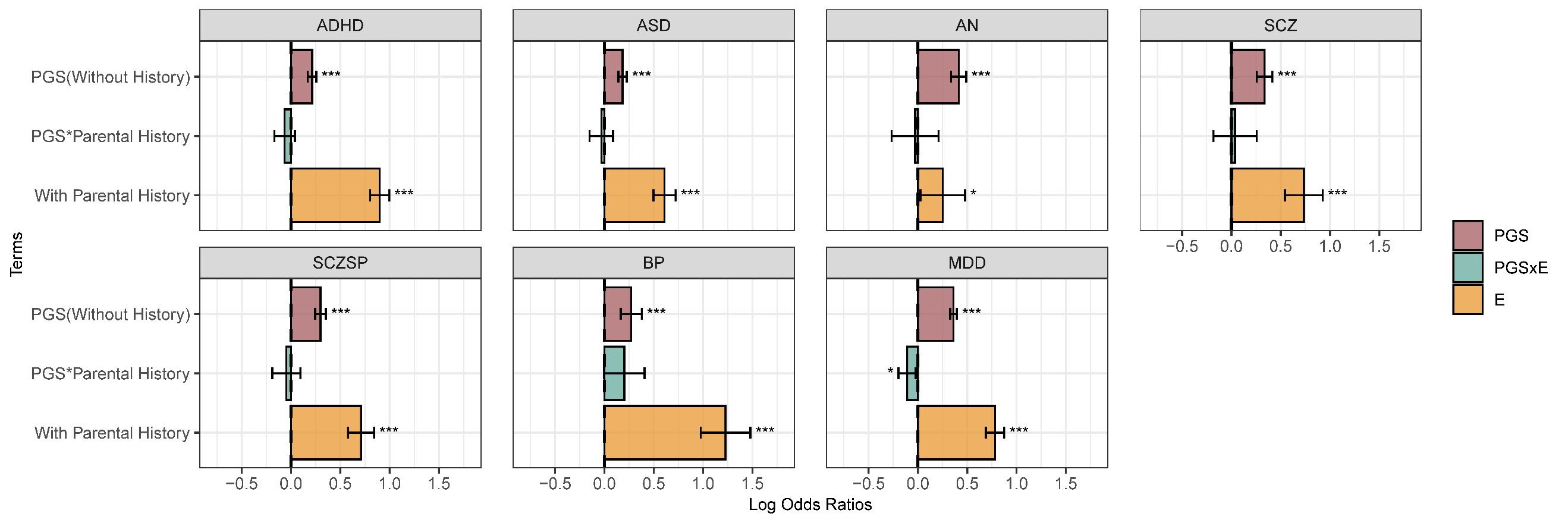

**S4 Figure. Parental history of psychiatric disorder analysis.** Log odds ratios and 95% confidence intervals are shown for each model term. No history of psychiatric disorder in any parent as the reference group. The **PGS** term represents the estimated effect of the polygenic score on the outcome within this reference category. The **PGS×E** (interaction) and E (environmental main effect) terms are interpreted relative to the reference category. Asterisks denote statistical significance at *p* < 0.05 *(), p < 0.01 (), and p < 0.001 ()*. AN: anorexia nervosa; SCZ: schizophrenia; SCZSP: Schizophrenia spectrum disorder; BP: Bipolar disorder.

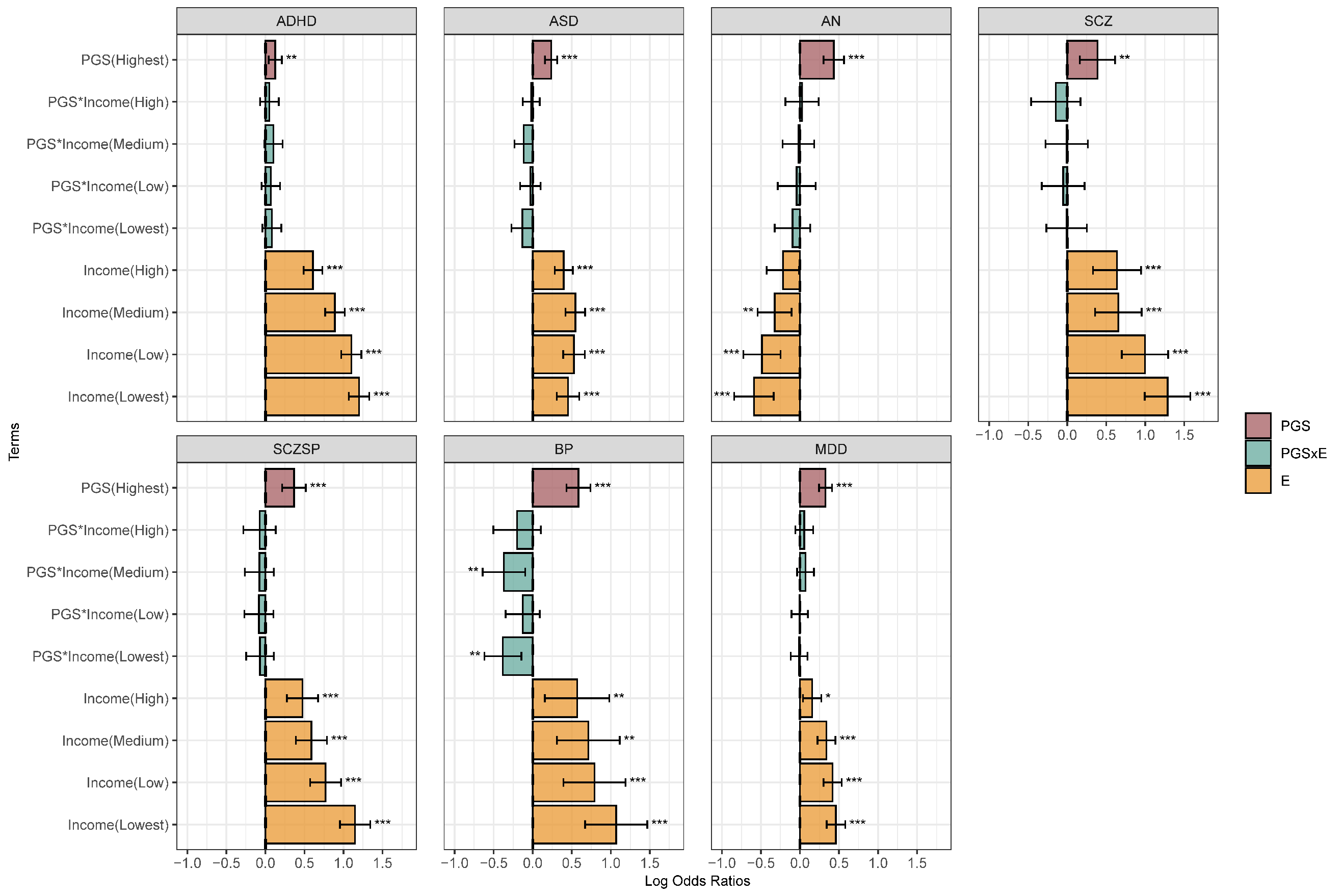

**S3 Figure. Parental income analysis.** Log odds ratios and 95% confidence intervals are shown for each model term. The top panel includes average parental income. The Very High category serves as the reference group. The PGS term represents the estimated effect of the polygenic score on the outcome within this reference category. The PGS×E (interaction) and E (environmental main effect) terms are interpreted relative to the reference category. Asterisks denote statistical significance at *p* < 0.05 *(), p < 0.01 (), and p < 0.001 ()*. AN: anorexia nervosa; SCZ: schizophrenia; SCZSP: Schizophrenia spectrum disorder; BP: Bipolar disorder.
